# Characterization of Ataxia-linked MFN2 Variants Reveals a Lipid Droplet/Mitochondrial Axis Driving Ferroptosis

**DOI:** 10.64898/2026.09.19.26362166

**Authors:** Mashiat Zaman, Armaan Mohan, Cole Chute, Fernanda Sousa Monteiro, Adriana Zardini-Buzatto, Gerald Pfeffer, Timothy E Shutt

## Abstract

Pathogenic variants in the mitochondrial protein Mitofusin 2 have long been recognized to cause the peripheral neuropathy Charcot-Marie-Tooth Type 2A (CMT2A). However, a small subset of MFN2 variants also impact the central nervous system and cause cerebellar ataxia. The reason for this discrepancy and the mechanisms underlying ataxia are unknown. Here we find a novel correlation between MFN2 variants linked to ataxia and increased susceptibility to ferroptosis. Mechanistically, we find that ataxia-linked MFN2 variants confer a gain-of-function that increases fatty acid import from lipid droplets into mitochondria, reshaping the cellular lipidome. This lipid redistribution primes cells for lipid peroxidation, ultimately increasing their susceptibility to ferroptotic cell death. Given that ferroptosis is linked to other ataxias (e.g., Friedreich’s ataxia), we propose that the increased sensitivity to ferroptosis within these MFN2 ataxia variants likely explains this specific phenotype. Thus, our findings uncover a gain-of-function for MFN2 ataxia variants that cause ferroptosis and likely contributes to cerebellar ataxia through a mechanism distinct from MFN2-mediated peripheral neuropathy.

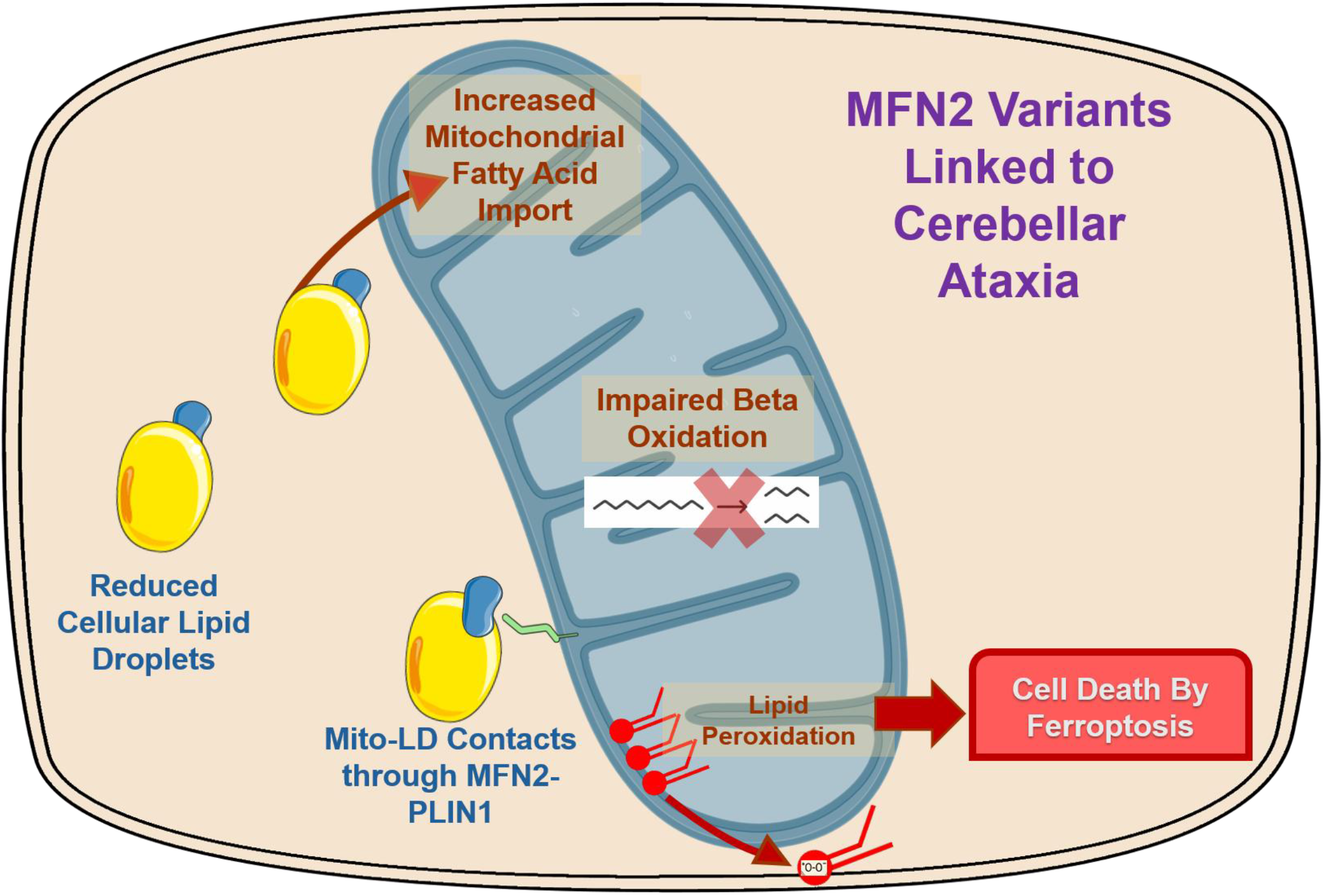
Graphical Abstract.

## Introduction

Pathogenic variants in the outer-mitochondrial-membrane (OMM) protein Mitofusin 2 (MFN2) are best known for causing the inheritable axonal peripheral neuropathy Charcot-Marie-Tooth disease subtype 2a (CMT2a). Although CMT2a is primarily considered a disease of the peripheral nervous system, a small subset of pathogenic MFN2 variants have been reported to cause cerebellar ataxia, which is an impairment of the central nervous system. While cerebellar involvement has been reported in some patients with CMT2a (Chen & Chan, 2009; Elbert et al., 2024; Lee et al., 2017; Madrid et al., 2020), a formal clinical diagnosis of ataxia remains uncommon in most cases. Moreover, recent studies have begun to recognize ataxia in patients with MFN2 variants who lack the peripheral neuropathy that is diagnostic of CMT2a (Elbert et al., 2024; Robert et al., 2025; Sharma et al., 2022). These observations raise the likelihood that ataxia represents a specific pathology with an underlying mechanism that is distinct from peripheral neuropathy.

A major challenge in understanding mechanisms of MFN2 disease is the multifunctional nature of MFN2, which was initially discovered as a mediator of mitochondrial outer membrane tethering and fusion (Chen et al., 2003). However, MFN2 is now recognized to perform several other functions, including mediating mitochondrial organelle contact sites (Boutant et al., 2017; de Brito & Scorrano, 2008; Hu et al., 2024), mitochondrial autophagy (Gegg et al., 2010) and mitochondrial motility (Misko et al., 2010). Among these diverse functions, MFN2 serves as a molecular tether at mitochondria-lipid droplet (mito-LD) contact sites (Bórquez et al., 2024; Boutant et al., 2017; Hu et al., 2024), which leads to a role in coordinating cellular lipid homeostasis (Boutant et al., 2017; Cai et al., 2024).

Lipid droplets are increasingly recognized as dynamic organelles that not only store excess lipids but also protect cells from lipotoxicity and oxidative damage. Consistent with a role for MFN2 in lipid homeostasis, loss of MFN2 results in an increased abundance of lipid droplets (Hu et al., 2024). Meanwhile, though only a small number of pathogenic MFN2 variants have been investigated for their impact on lipid droplets, for the most part they also cause increased abundance (Zaman & Shutt, 2022). However, we recently reported on the ataxia-associated MFN2 variant D414V, which displayed the opposite phenotype of decreased lipid droplet abundance (Sharma et al., 2022). This observation raised the possibility that this specific lipid perturbation may be relevant to the ataxia described in the patient.

An emerging theme across inherited cerebellar ataxias is disruption of lipid homeostasis (Zhao et al., 2022). Pathogenic variants in several lipid homeostasis genes, including *SNX14*, *PNPLA6*, *ABHD12* and *ELOBL4/5*, cause cerebellar dysfunction (Datta et al., 2019; Zhao et al., 2022). A key consequence of disrupted lipid homeostasis is increased vulnerability to lipid peroxidation and ferroptosis (Yang & Stockwell, 2016), a form of regulated cell death by oxidation of polyunsaturated phospholipids. Consistent with this notion, ferroptosis has been increasingly implicated in cerebellar dysfunction and ataxias (La Rosa et al., 2020; Turchi et al., 2020). Lipid droplets have been shown to protect against lipid peroxidation through sequestering excess fatty acids and peroxidation-prone lipids away from cellular membranes (Sun et al., 2025). Despite growing evidence linking both MFN2 and ataxias to lipid biology, it remains unknown whether lipid homeostasis contributes to MFN2-associated cerebellar disease.

We therefore hypothesized that dysregulated lipid homeostasis represents a defining feature of MFN2-associated ataxia. To test this idea, we examined lipid metabolism across a panel of pathogenic MFN2 variants expressed in an isogenic background, as well as in patient-derived cells. Amongst cells expressing ataxia-linked MFN2 variants, we identify profound differences in lipid homeostasis and uncover a link to lipid peroxidation and ferroptosis susceptibility. These findings provide insight into the pathological heterogeneity connected to MFN2 dysfunction by revealing a mechanistic distinction between MFN2-associated ataxia and MFN2-linked peripheral neuropathy.

## Methods

### Cell Culture

U2OS MFN2 KO cells were a generous gift from Dr. Edward Fon (McGill University, Canada). U2OS cells were maintained in Dulbecco’s Modified Eagle Medium (Thermo Fisher Scientific, 11965118), supplemented with 10% fetal bovine serum (FBS) (Fisher Scientific, 12483020), and incubated at 37 °C and 5% CO_2_. Fibroblasts used in the study have been reported on before: R334K (https://doi.org/10.1101/2024.09.05.24313021), Q367H (Zaman et al., 2025) and R707W (Sawyer et al., 2015). The Conjoint Health Research Ethics Board of the University of Calgary gave ethical approval for the use of patient fibroblasts in this work. Fibroblasts were cultured in Minimum Essential Medium (Thermo Scientific, 11095098), supplemented with 10% FBS, and maintained at 37 °C and 5% CO_2_.

### Transduction

Selected MFN2 variants of interest were introduced through viral transduction. Phoenix cells (ATCC, CRL-3213) were transfected with a retroviral vector including the MFN2 open reading frame and a mitochondria-targeted mNeonGreen sequence, using Lipofectamine 3000 (Thermo Fisher Scientific, L3000001). Virus was collected for three consecutive days, concentrated with Lenti-X Concentrator (Takara Bio, 631232) and added to U2OS MFN2 KO cells for 72 hours. Transduced cells were then sorted for mNeonGreen expression on a SONY SH800 cell sorter (Flow Cytometry Facility, University of Calgary). Expression levels of WT MFN2/variants were verified using western blot analysis.

### Western Blot

Western blot analyses were performed as previously described (https://doi.org/10.1101/2022.10.03.510695). Briefly, cells were trypsinized (VWR, CA45000-664) and total protein lysate was collected using RIPA lysis and extraction buffer (Thermo Fisher Scientific, 89901), supplemented with protease inhibitor (1:100) (VWR, 97063-010). Protein levels were quantified through a BCA assay (Thermo Fisher Scientific, 23223/23224) using BSA standards (Thermo Scientific, 23235) and 30 µg protein was loaded into each well of an SDS-PAGE gel. Following overnight transfer onto a PVDF membrane (Bio-Rad, 1620177), the blots were probed for MFN2 and Alpha-Tubulin. Primary antibodies used were anti-MFN2 (Santa Cruz Biotechnology, sc-515647) and anti-Alpha-Tubulin (DSHB, 12G10). The secondary antibody used was goat anti-Mouse HRP (Thermo Scientific, 31430). The blots were subsequently visualized using Femto Chemiluminescent Substrate (Fisher Scientific, PI34095), using a Bio-Rad imager.

### Live Cell Imaging

Confocal live cell imaging was performed using the same system previously described (Bora et al., 2025). Briefly, 35,000 cells were grown for 24 hours in live cell imaging dishes (Cellvis, D35-20-1.5-N), before being imaged on an Olympus Spinning Disk Confocal System (Olympus SD-OSR) was used with a 60× oil immersion objective (UPlanApo 1.50 Oil HR/0.13-0.19), 100x oil immersion objective (UApo 100x/1.49/0.13-0.19) or a 40x objective (UPlanSApo/40x/0.95/0.11-0.23). Neutral lipid droplets were imaged using LipidSpot 610 (Biotium, 70069) and mitochondria were visualized with MitoView Blue (Cedarlane Labs, 70070BT). Fluorescent fatty acids were visualized using BODIPY 558/568 C_12_ (Fisher Scientific, D3835) and mitochondria were labelled in the same experiment with MitoTracker Deep Red (Thermo Fisher Scientific, M22426). β-oxidation was quantified using FAO Blue fatty acid oxidation detection reagent (DiagnoCine, FNK-FDV-0033). Total cellular lipid peroxidation levels was analyzed using BODIPY 665/676 lipid peroxidation sensor (Thermo Scientific, B3932) and mitochondrial lipid peroxidation was analyzed using OxiVision Red Mitochondrial Lipid Peroxidation Sensor (Cedarlane Labs 21510AAT). Cell death was analyzed using SYTOX Orange Nucleic Acid Stain (Thermo Scientific, S11368). Glutathione levels were analyzed using Thioltracker™ Violet glutathione detection reagent (Thermo Scientific, T10095), iron levels were detected using FerroOrange (New England Biolabs, 36104S) and mitochondrial superoxides were analyzed using MitoSOX (Thermo Scientific, M36008).

### Image Analysis/Quantification

Lipid droplet number and size were quantified using the ‘Analyze Particles’ plugin on FIJI (Schindelin et al., 2012). Colocalization analyses where we quantified Pearson Colocalization Coefficients for mitochondrial fatty acid import were performed using JaCoP on FIJI (Bolte & Cordelières, 2006). The levels of fluorescence intensity for quantifying β-oxidation, mitochondrial lipid peroxidation and total lipid peroxidation were performed using the ‘Measure’ plugin on FIJI (Schindelin et al., 2012).

### Drug Treatments

The inducers of ferroptosis used in this study were used at the following doses: H_2_O_2_-500µM (Sigma-Aldrich, 216763), Ferrostatin-1-1µM (MedChem Express, HY-100579), Erastin-5µM (MedChem Express, HY-15763) and RSL3-1µM (MedChem Express, HY-100218A). All drug treatment times are mentioned in the respective figures.

### Lipidomics

Lipidomics data were acquired at the Calgary Metabolomics Research Facility (CMRF) at the University of Calgary. Lipidomics analyses were performed in accordance with established protocols (Zardini Buzatto et al., 2020). Briefly, one aliquot of each sample (in triplicate) was extracted with a modified Folch method (dichloromethane/ methanol 2:1)1-4 and Avanti Splash Lipidomix Mass Spec standard mixture (14 deuterated lipids). A pool was prepared with a small aliquot of the organic phase from each sample for quality control. An aliquot of the organic layer was evaporated to dryness using a SpeedVac and resuspended in chromatographic mobile phases. Samples were then analyzed by reversed-phase LC-MS/MS using a C18 column in a Thermo Vanquish LC coupled to a Thermo Q-Exactive HF Quadrupole-Orbitrap mass spectrometer. The chromatograms were processed using in-house developed software (peak picking, alignment, data cleansing, annotations using MS/MS spectral match and mass-match, batch correction, and normalization by internal standards and to the median), developed by the Buzatto Research Group at the University of Calgary.

Lipid species were annotated in accordance with established protocols (Liebisch et al., 2020). High-confidence annotations were based on MS/MS fragmentation patterns, with a match score of 400 or higher (on a 0–1000 scale). Low-confidence annotations were made by matching the mass-to-charge (m/z) ratio of the compound to lipid databases within 3.0 ppm or 3.0 mDa. All annotations are filtered using expected retention times, ionization patterns, and known biological context. The heatmaps generated represent the most important lipids for group separation, selected by one-way ANOVA. In the heatmap, higher peak intensities (i.e., greater relative abundances) are shown in red, and lower abundances are shown in blue, allowing visual comparison across samples.

We assessed dataset normality using the Shapiro–Wilk test and homoscedasticity using Levene’s test. If more than 30% of annotated lipids violate the assumptions for parametric testing, a non-parametric test (Kruskal–Wallis, Mann-Whitney U test) is applied as needed. We assessed dataset normality using the Shapiro–Wilk test and homoscedasticity using Levene’s test. If more than 30% of annotated lipids violated the assumptions for parametric testing, a non-parametric test (Kruskal–Wallis, Mann-Whitney U test) was applied as required.

### Plate Reader Assays

Fluorescent plate reader assays were used to quantify fluorescence for the cell death assays, iron levels, GSH levels and mitochondrial superoxide levels. For the plate reader assays, 10,000 cells were seeded into each well of 96-well cell culture plates (MaxBioChem, MBC.TC.F. 96) and grown for 24 hours. Following dye incubation and washes with media, fluorescence was measured using a SpectraMax i3x plate reader (Molecular Devices) with incubation at 37 degrees during the reading phase, connected to SoftMaxPro 6 software. Fluorescence intensity readings were calibrated against empty wells with no fluorescent dyes, and data were normalized by quantifying protein levels in each well using BCA assays, normalized to BSA.

## Results

### MFN2 variants from patients with ataxia show reduced lipid droplets

While only a handful of MFN2 variants have been studied for their effects on lipid droplets, most of the studied variants lead to an increase in lipid droplet size and abundance. However, reduced size and abundance of lipid droplets was reported for the MFN2 D414V variant (Sharma et al., 2022). Given this previous work was done by different groups, to revisit this discrepancy, we performed a direct comparison of lipid droplets in fibroblasts from patients with distinct MFN2 variants to which we had access (R334K/Q367H/D414V/R707W). The R334K variant represents one of the most severe cases of MFN2 disease, with patient cells exhibiting global and severe MFN2 dysfunction (Zaman et al., 2026). Meanwhile, the MFN2 Q367H variant also represent a distinct pathology from CMT2A, with mtDNA release and inflammation in patient cells underlying myopathy. Finally, the R707W variant has been linked to CMT2A when heterozygous, as well as multiple symmetric lipomatosis when homozygous (Sawyer et al., 2015), making it an intriguing candidate with respect to lipid metabolism. Live cell imaging with LipidSpot 610 confirmed previous findings, as only the MFN2 D414V patient fibroblasts showed a reduction in lipid droplets. Meanwhile, the other MFN2 variant fibroblasts (R334K, Q367H and R707W) showed an increase in lipid droplet abundance and size (Extended Figure 1A a-c), similar to what has been reported with MFN2 knockout (Cai et al., 2024; Hu et al., 2024; Mancini et al., 2019). Therefore, while alterations to lipid droplets are a consistent consequence of pathogenic MFN2 variants, the direction of these changes depends on the variants. In this regard, the increased lipid droplet size and abundance in R334K, Q367H and R707W variants is likely due to a loss-of-function. Conversely, the reduced lipid droplet size and numbers seen with the D414V variant appears to be due to a gain-of-function.

To expand the number of MFN2 variants and eliminate confounding effects of variable genetic backgrounds in patient fibroblasts, we employed a knockout re-expression approach where wildtype (WT) MFN2 or selected variants of interest were re-expressed in MFN2 KO cells (McLelland et al., 2018) (Figure 1a-b/Table1). The selected MFN2 variants, including those from the fibroblast studies above, were chosen for several reasons. The R94Q, R259C, and R364W variants were chosen as they cause classic CMT2A, with R94Q and R364W the most frequently reported and studied MFN2 variants. We chose T105M, T206I, R280C, and D414V variants, which are all linked to cerebellar ataxia, to further explore the mechanisms underlying this pathology. Finally, the remaining MFN2 variants (D210V/R334K/Q367H/R400Q/L643P/R707W) are linked to a variety of pathogenic phenotypes (Table 1).

**Figure 1:**
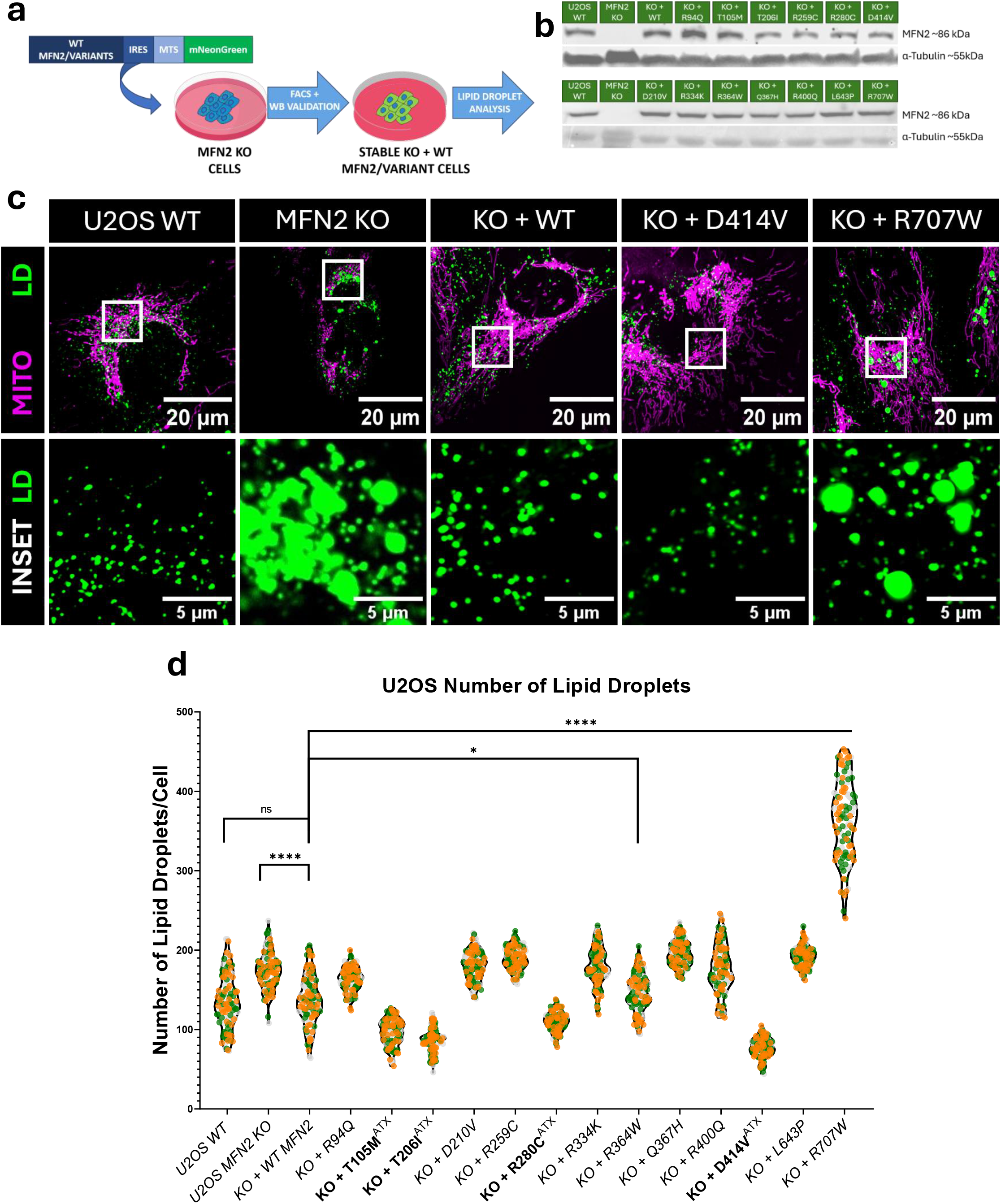
Ataxia-linked MFN2 variants show reduced cellular lipid droplets. (a) Schematic representation of the approach for re-expressing selected MFN2 variants in U2OS MFN2 KO cells and (b) western blot validation of re-expression of selected variants in KO cells. (c) Representative live-cell imaging confocal images (100x objective) showing lipid droplets (labelled with LipidSpot 610) and the mitochondrial network (labelled with MitoTracker Green). The white boxes in the top panels represent the area zoomed into in the bottom panel. (d) Quantitative analysis of the number of lipid droplets per cell in U2OS WT/MFN2 KO or re-expression variants; the ataxia-specific variants are indicated on the graph. The violin plots show the median and interquartile range; each dot represents a cell, and colours represent biological replicates. The statistical symbols on the graphs indicate: ns P > 0.05, * P ≤ 0.05, **** P ≤ 0.0001.

**Table 1:**
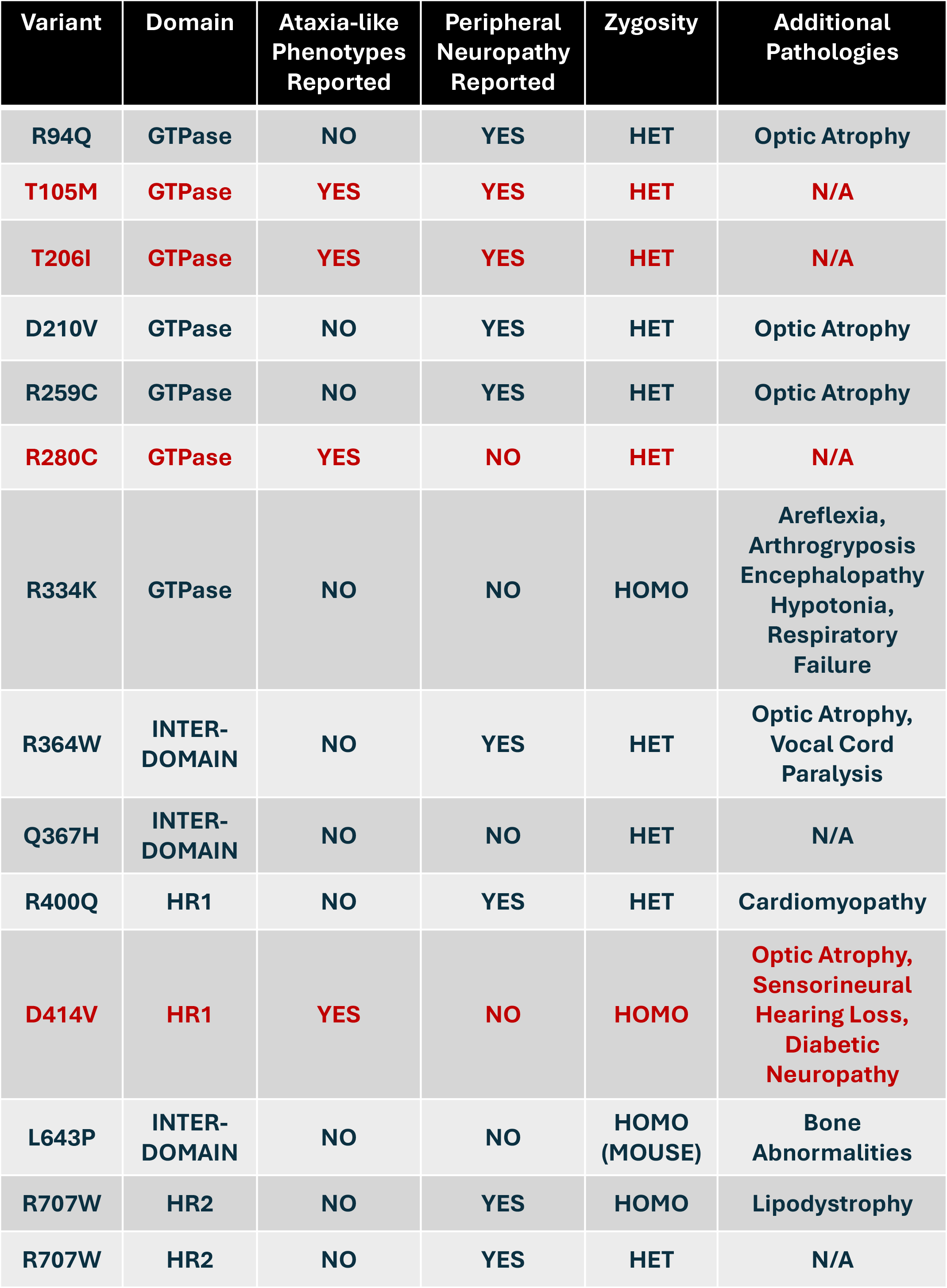
List of MFN2 Variants Studied. The table lists all the MFN2 variants studied here, with the cerebellar ataxia-linked variants indicated in red. The table also lists the respective domains, reports of peripheral neuropathy, patient genotypes, and any additional pathologies.

With this panel of cells established, we next investigated whether and how these MFN2 variants affect lipid droplets. Of our panel of fourteen variants (including WT re-expression), the ataxia-linked variants (T105M/T206I/R280C/D414V) all showed a decrease in lipid droplet number (Figure 1c-d, Extended Figure 1B a), mirroring findings in patient fibroblasts. There was also a general decrease in the size of lipid droplets in the ataxia variants, except for T105M, which had larger lipid droplets (Extended Figure 1B a-b). Meanwhile, the general trend for the remaining MFN2 variants was an increase in the size and number of lipid droplets, which was also observed in MFN2 KO cells, except for MFN2 R364W, which resembled WT in terms of LD size (Figure 1c-d, Extended Figure 1B a-b). Overall, most MFN2 variants displayed a lipid droplet phenotype of increased abundance that is consistent with a loss-of-function seen in MFN2 KO cells. However, MFN2 variants from patients with an ataxia phenotype showed the opposite trend with reduced lipid droplet abundance.

### Ataxia-linked variants show increased mitochondrial fatty acid import

To simplify subsequent mechanistic investigations, we reduced our pool of re-expression variants. In addition to all of the ataxia variants (T105M, T206I, R280C, and D414V), we included R94Q (the most well-studied and abundant MFN2 variant found in CMT2a patients), R334K (a novel variant causing a severe neonatal fatal onset disorder (Zaman et al., 2026), Q367H (linked to myopathy), (Zaman et al., 2025), and R707W, a variant linked to lipodystrophy (Sawyer et al., 2015).

To investigate why lipid droplet abundance was altered in this panel of MFN2 variant lines, we examined the transfer of lipids from lipid droplets to mitochondria using BODIPY C12 558/568. This fluorescently labelled fatty acid analogue is readily imported into lipid droplets with a 16-hour pulse and is typically then transferred to mitochondria with a 24-hour chase under glucose-starvation conditions (Miner & Cohen, 2024) (Figure 2a). Using this assay, there is a significant reduction in fatty acid transfer into mitochondria in cells with larger and more abundant lipid droplets (e.g., MFN2 KO cells and cells re-expressing variants R94Q, R334K, Q367H and R707W)(Fig 2C). This finding suggests that increased lipid droplets in these lines results from reduced lipid flux from lipid droplets to mitochondria. In contrast, while the ataxia variants behaved similarly to WT MFN2 under starvation conditions that induce lipid droplet-to-mitochondrial lipid transfer, they displayed increased fatty acid import into mitochondria when grown in standard glucose media (Figure 2b-c). This finding suggests that an upregulation of mitochondrial fatty acid import from lipid droplets explains the reduction in cellular lipid droplets in the ataxia-linked variants.

**Figure 2:**
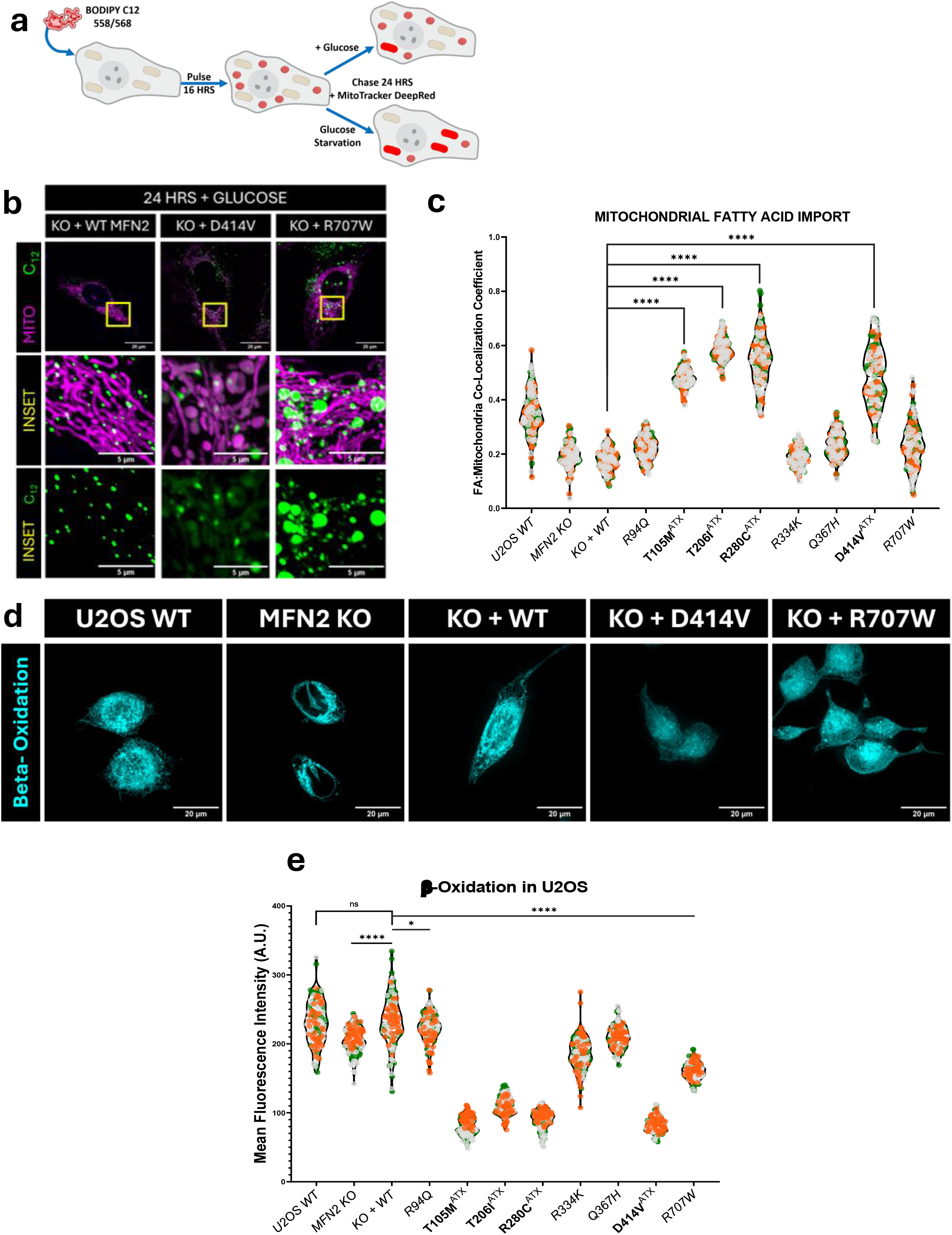
Ataxia linked MFN2 variants show increased mitochondrial fatty acid import but reduced β-oxidation. (a) Schematic representation of fatty acid pulse-chase assay, where cells of interest are pulsed with BODIPY 558/568 for 16 hours, incubated with Mitotracker DeepRed, then chased with or without glucose present in the media for 24 hours. (b) Representative confocal live cell images (60x objective) showing mitochondria (MitoTracker DeepRed) and fatty acids (BODIPY 558/568) in U2OS cells after chasing with glucose supplementation for 24 hours. The yellow boxes in the top panels represent the area zoomed into in the bottom panel. (c) Quantitative analyses of the Pearson Co-localization coefficient of mitochondria and BODIPY 558/568 fatty acids in U2OS cells. (d) Representative confocal live cell images (60x objective) of U2OS cells with FAOBlue, incubated without glucose for 24 hours. (e) Quantitative analyses of fluorescence intensity indicative of β-oxidation in U2OS cells under glucose starvation for 24 hours; the ataxia-specific variants are indicated on the graph. The violin plots in (c) and (e) show median and interquartile range, each dot represents a cell and colours represent biological replicates. The statistical symbols on the graphs indicate: ns P > 0.05, * P ≤ 0.05, **** P ≤ 0.0001.

### Ataxia-linked variants show impaired β-Oxidation

To investigate what happens to the fatty acids imported into mitochondria from lipid droplets, we investigated β-oxidation, which is the typical fate of these lipids (Clarke, 1990). We evaluated β-oxidation using the mitochondrial fatty acid oxidation sensor (FAOBlue), which fluoresces upon oxidation of the dye. While MFN2 KO cells and non-ataxia MFN2 variant lines (R94Q/R334K/Q367H/R707W) had a slight reduction in β-oxidation, unexpectedly, despite higher fatty acid import, the ataxia-linked variants had the lowest levels of β-oxidation (Figure 2d-e).

### Ataxia-linked MFN2 D414V variant exhibits distinct lipidome remodelling

Given the lipid alterations observed in the ataxia-specific variants, we next investigated changes in the whole-cell lipidome. Here, we used a further reduced subset of KO-re-expression cell lines, including the MFN2 D414V ataxia variant and the MFN2 R707W variant. Notably, re-expression of WT MFN2 in KO cells restored the lipidome to levels similar to WT cells (Figure 3A). However, we see a distinct set of lipidome perturbations in cells re-expressing the D414V variant compared with all other groups (Figure 3A), particularly compared with WT re-expression (Figure 3B). Compared to cells re-expressing WT MFN2, the cells re-expressing the MFN2 D414V ataxia variant had 219 lipid species that were significantly increased and 276 lipid species that were significantly decreased (Figure 3b). As a control, when comparing lipid abundance in the KO cells re-expressing WT MFN2 to the parental U2OS cells, there were only 13 lipid species that were significantly increased, and 4 species significantly decreased (Extended Figures 3b, 3d). This finding suggests that re-expression of the full-length WT MFN2 restores most of the lipid perturbations caused by loss of MFN2. Among the prominent changes in D414V cells, we observed increases in phosphatidylethanolamine (PE), phosphatidylcholine (PC), and sphingomyelins (SM) (Figure 3a), along with a global reduction in TAG levels (Figure 3c/Extended figure 3 g-h).

**Figure 3:**
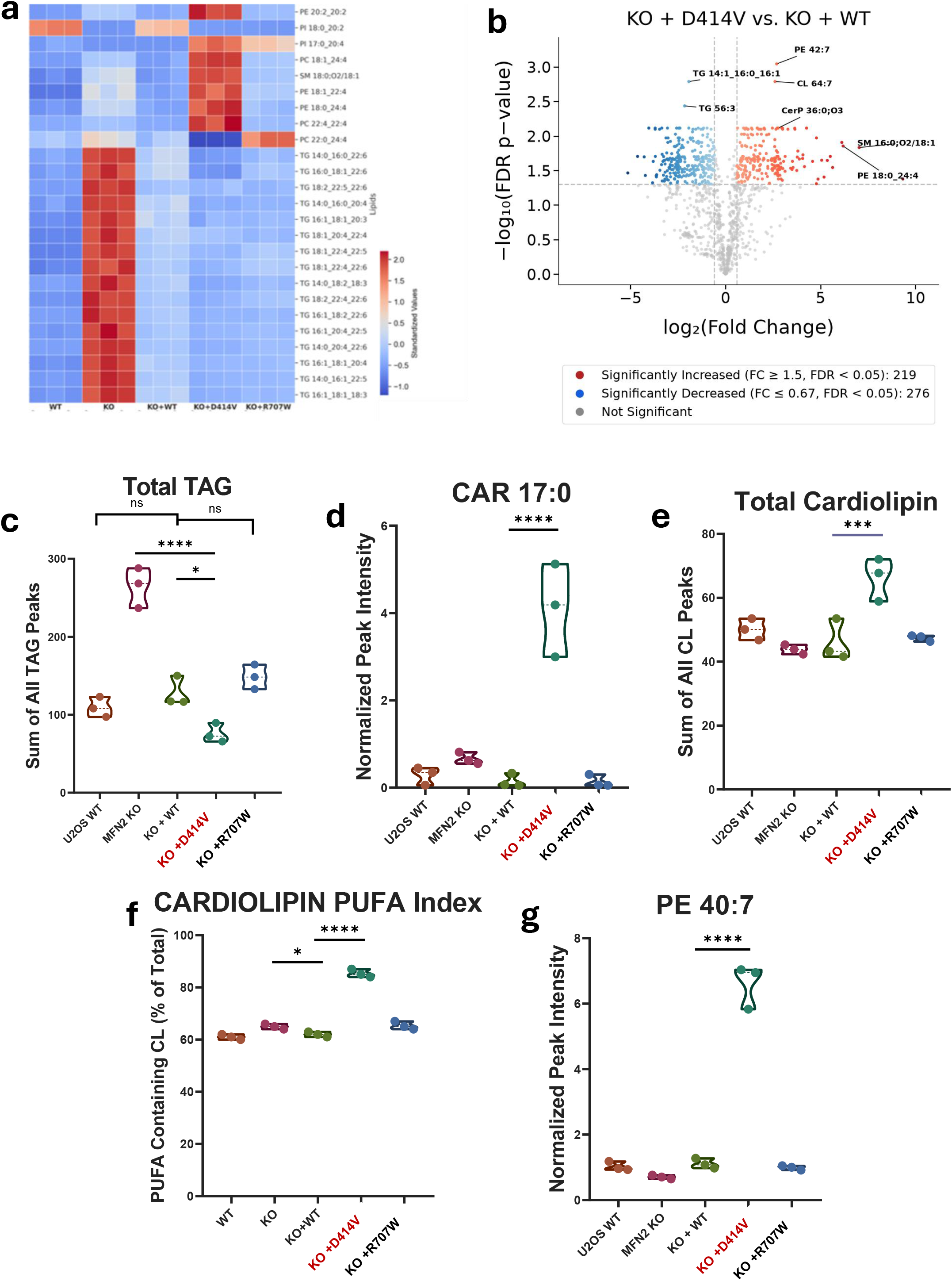
Remodelling of the lipidome in MFN2 D414V re-expression cells. (a) Representative heatmap showing the top 25 upregulated and downregulated lipid species in U2OS cells. (b) Representative volcano plot comparing U2OS MFN2 KO re-expressing D414V against KO cells re-expressing WT MFN2. The top significant species are labelled on the plot. FC: fold-change for Group A / Group B. FDR-p: p-value corrected for false discovery rate (Benjamini-Hochberg correction). Lipids are considered significantly altered for FC ≥1.50 or ≤0.667, FDR-corrected p-value < 0.05. (c-e) Quantitative analyses of the (c) sum of TAG species, (d) levels of Acylcarnitine 17:0 species and (e) sum of all cardiolipin species in U2OS cells. (f) Quantitative analyses of unsaturated cardiolipin species as a percentage of total cardiolipin species in U2OS cells. (g) Quantitative analyses phosphatidylethanolamine 40:7 levels in U2OS cells. The violin plots in (c-g) indicate the median and IQR of each analysis, n=3 biological replicates.

Notably, two findings in the lipidomic profiling support earlier observations in cells with MFN2 ataxia variants. First, D414V cells showed a reduction in the total triacylglycerol (TAG) levels (Fig 3c), the main substance found in lipid droplets (Fujimoto & Parton, 2011), which is consistent with the reduced lipid droplets observed with ataxia variants. Second, D414V cells had an accumulation of acylcarnitine species (Cortassa et al., 2017; Guerra et al., 2022) (Figure 3d/Extended Figure 3f), which is consistent with incomplete β-oxidation observed in the ataxia variants.

Given MFN2 is associated with mitochondrial functions, we also investigated the mitochondrial-specific lipid cardiolipin. Again, showing a unique phenotype compared to the other cell lines, D414V cells exhibited an ∼40% increase in the total cardiolipin content (Figure 3e), which was also enriched in polyunsaturated fatty acid (PUFA) side chains (Figure 3f/Extended Figure 3h-i).

### Increased lipid peroxidation in ataxia-linked MFN2 variants

Given that PUFAs are susceptible to lipid oxidation that can be detrimental (Ayala et al., 2014; Hill et al., 2012; Mortensen et al., 2023; Pope & Dixon, 2023), we investigated global lipid peroxidation in our secondary panel of MFN2 KO cells re-expressing selected MFN2 variants using the BODIPY 665/676 dye ratio. Notably, following hydrogen peroxide (H_2_O_2_) stress, the ataxia-linked variants had increased peroxidized lipids (Figure 4a), leading to an approximate 5-fold increase in the ratio of peroxidized to non-peroxidized signal relative to control (Figure 4b). Given that the lipid peroxidation signal in the ataxia variants appeared to colocalize with the mitochondrial network (Figure 4a), we then examined mitochondrial lipid peroxidation using the mitochondrial-specific lipid peroxidation sensor OxiVision Red. Here, we observed a ∼4-fold increase in mitochondrial peroxidation levels in the ataxia-linked variants (Figure 4c-d).

**Figure 4:**
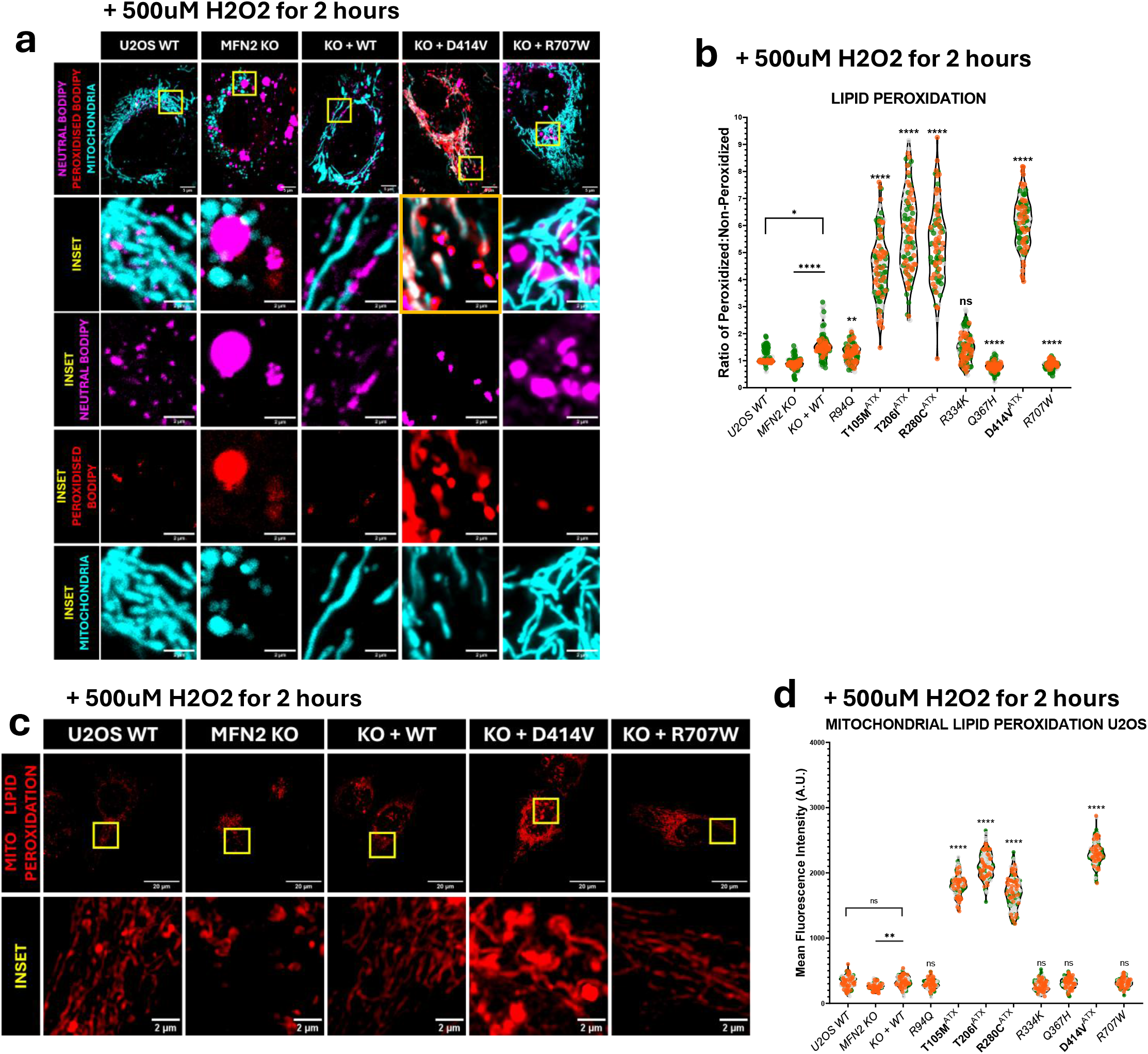
Increased lipid peroxidation in ataxia-linked variants. (a) Representative confocal live cell images (60x objective) of mitochondria (MitoView Blue) and lipid peroxidation (BODIPY 665/676), with 500µM hydrogen peroxide added for 2 hours. The magenta signal indicates non-oxidized lipids and red indicates the peroxidized lipids. The yellow boxes in the top panels represent the area zoomed into in the bottom panel. The image outlined with the yellow box shows co-localization between the mitochondrial network and peroxidized lipids. (b) Quantitative analysis of the ratio of peroxidized to neutral lipids in U2OS cells with 500µM hydrogen peroxide added for 2 hours. (c) Representative confocal live cell images (60x objective) showing mitochondrial lipid peroxidation with 500uM hydrogen peroxide added for 2 hours, measured with OxiVision Red. (d) Quantification of mitochondrial lipid peroxidation with 500µM hydrogen peroxide added for 2 hours in U2OS cells. The violin plots in (b) and (d) indicate the median and IQR; each dot represents a cell and colours represent biological replicates. The statistical symbols on the graphs indicate: ns P > 0.05, * P ≤ 0.05, ** P ≤ 0.01, **** P ≤ 0.0001.

### Increased ferroptotic cell death in ataxia-linked MFN2 variants

Excessive lipid peroxidation can promote ferroptosis, a caspase-independent cell death pathway reliant on iron-mediated phospholipid lipid peroxidation, which is linked to ataxia. Thus, we investigated ferroptotic cell death in our MFN2 variant lines. To this end, following H_2_O_2_ treatment as a general inducer of oxidative stress, we measured cell death using the Sytox orange dye, which labels dying cells (Figure 5a-b). As expected, the ataxia-linked variants consistently had the highest levels of cell death compared to the other MFN2 variants (Figure 5b-c). Importantly, we confirmed that this cell death occurs through ferroptosis, as the selective ferroptosis inhibitor ferrostatin-1 (FS-1) completely prevents the cell death in all the cell lines (Figure 5c). To further interrogate ferroptosis in these cell lines, they were treated with Erastin and RSL3, two specific inducers of ferroptosis that function through distinct mechanisms: erastin inhibits the cystine/glutamate antiporter, while RSL3 the inactivates glutathione peroxidase 4 (GPX4). Again, the ataxia variants showed increased susceptibility to both ferroptosis-inducing treatments (Extended Figure 5b-c), which was rescued by co-treatment with FS-1.

**Figure 5:**
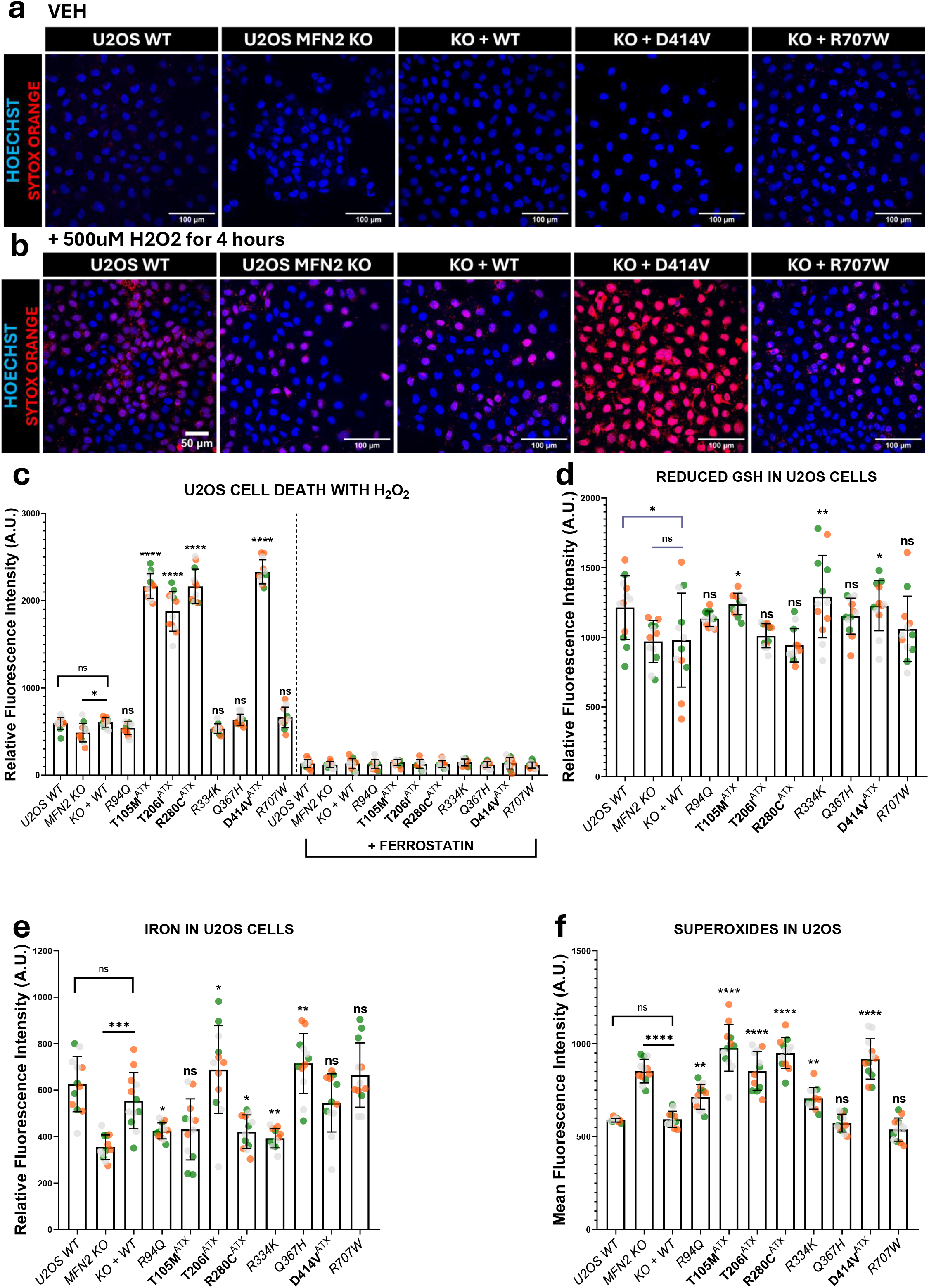
Increased ferroptotic cell death in ataxia-linked variants. (a-b) Representative confocal live cell images (40x objective) (a) without treatment and (b) with 500µM hydrogen peroxide for 4 hours showing all nuclei (Hoechst) and dead cells (red). (c) Quantitative analysis of fluorescent intensity from Sytox Orange dye indicating cell death in U2OS cells, treated with 500uM hydrogen peroxide for 4 hours or with hydrogen peroxide and 1µM ferrostatin. (d-f) Quantitative analyses of (d) levels of reduced GSH, (e) levels of iron and (f) levels of mitochondrial superoxides in U2OS cells. The bars and lines in graphs (c-f) show the mean +/-SD, each point indicates a well and colours represent biological replicates. The statistical symbols on the graphs indicate: ns P > 0.05, * P ≤ 0.05, ** P ≤ 0.01, *** P ≤ 0.001, **** P ≤ 0.0001.

Next, we examined key redox players involved in ferroptosis, including GSH levels, which quench lipid peroxidation; iron levels, which drive lipid radical production through the Fenton reaction; and mitochondrial superoxides, which cause lipid peroxidation. Although there were some fluctuations among the cell lines, we did not observe any trends in GSH levels (Figure 5d) or iron (Figure 5e) that correlated specifically with the ataxia variants. However, we did observe a general increase in mitochondrial superoxide levels in the ataxia-linked variants, which were consistently higher than in any of the other MFN2 variants (Figure 5f). Combined with the altered lipids in cells with ataxia variants described above, our findings suggest that the altered lipid composition and increased superoxide production, rather than levels of reduced GSH or increases in iron, make the cells expressing ataxia variants more susceptible to lipid peroxidation and ferroptosis.

### Fibroblasts from the D414V ataxia patient show lipid alterations and ferroptotic cell death

Lastly, we set out to confirm the mechanistic findings from U2OS cells in the primary MFN2 patient fibroblasts to which we had access, including the D414V ataxia variant. Starting with the lipid phenotype, as in the U2OS ataxia variant cells, the D414V fibroblasts had increased mitochondrial fatty acid import under glucose supplementation (Figure 6a-b, Extended Figure 6A a) and decreased β-oxidation relative to the control and other MFN2 variant fibroblast lines (Figure 6c). We also observed significant alterations to the lipidome of D414V patient fibroblasts compared to control fibroblasts, with a significantly increased abundance of 233 species and decreased levels of 316 species (Extended Figure 6Ba). Notably, the changes in the D414V fibroblast lipidome were consistent with the findings in the D414V U2OS cells, including a decrease in TAG species (Extended Figure 6B b-c), an accumulation of acylcarnitine species (Extended Figure 6B d), and an increase in PUFA species (Extended Figure 6B b/e). Collectively, the shared lipid changes in U2OS cells re-expressing MFN2 ataxia variants and D414V patient fibroblasts show that MFN2 ataxia variants cause a conserved cellular phenotype with reduced lipid droplets, increased fatty acid import from lipid droplets to mitochondria, impaired β-oxidation, and a buildup of PUFA-containing lipids.

**Figure 6:**
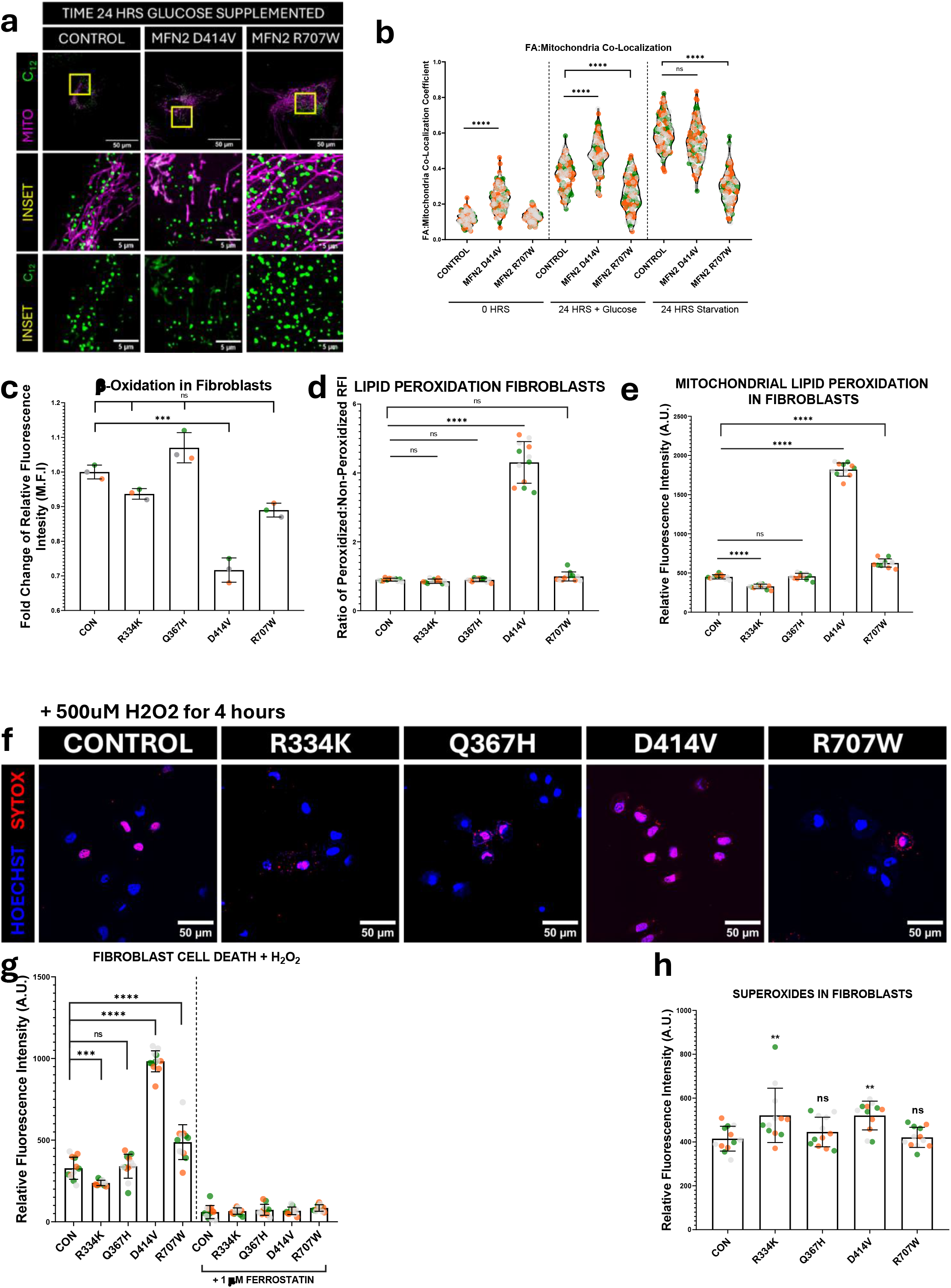
Ataxia-linked fibroblast replicates ferroptotic phenotypes. (a) Representative confocal live-cell images (60x objective) showing mitochondria (MitoTracker Green) and fatty acids (BODIPY 558/568) in healthy and MFN2 patient fibroblasts supplemented with glucose and chased for 24 hours. The yellow boxes in the top panels represent the area zoomed into in the bottom panel. (b) Quantitative analyses of the Pearson Co-localization coefficient of mitochondria and BODIPY 558/568 fatty acids in fibroblasts. The violin plots in (b) indicate the median and IQR, each dot represents a cell and colours represent biological replicates. (c) Quantification of β-oxidation using FAOBlue dye in fibroblasts. (d) Quantification of the ratio of peroxidized lipids to neutral lipids using BODIPY 665/676 in fibroblasts. (e) Quantification of mitochondrial lipid peroxidation using OxiVision Red Mitochondrial Lipid Peroxidation Sensor in fibroblasts. (f) Representative confocal live cell images (40x objective) with 500µM hydrogen peroxide for 4 hours showing all nuclei (Hoechst) and dead cells (red) in fibroblasts. (g) Quantification of fluorescent intensity from Sytox Orange dye indicating cell death in fibroblasts, treated with 500µM hydrogen peroxide for 4 hours or with hydrogen peroxide and 1µM ferrostatin. (h) Quantification of levels of mitochondrial superoxides in fibroblasts. The bars and lines in (c-e/g-h) indicate mean +/-SD, each point indicates a well and colours represent biological replicates. The statistical symbols on the graphs indicate: ns P > 0.05, ** P ≤ 0.01, *** P ≤ 0.001, **** P ≤ 0.0001.

We also investigated the consequence of these lipid alterations on ferroptosis in patient fibroblasts. Again, as with the U2OS cells expressing MFN2 ataxia variants, the D414V fibroblasts showed increased lipid peroxidation relative to the other fibroblast lines both globally (Figure 6d) and in mitochondria (Figure 6e). Relative to control fibroblasts, the D414V fibroblasts also exhibited increased ferroptotic cell death in response to H_2_O_2_, Erastin, and RSL3 (Figure 6f-g/Extended Figure 6b-d) and increased levels of mitochondrial superoxide (Figure 6h), with no significant changes in the levels of GSH or iron (Extended Figure 6e-f). Thus, we see similar trends in the D414V patient fibroblasts as in the U2OS MFN2-KO cells re-expressing D414V and other ataxia variants. The conservation of these findings across two distinct cellular models supports a novel disease mechanism in which MFN2 ataxia variants alter lipid homeostasis, promoting susceptibility to ferroptosis.

## Discussion

Although MFN2-associated disease is commonly studied in the context of peripheral neuropathy, a small subset of MFN2 variants is linked to ataxia. Here, we identify a conserved metabolic signature linked to MFN2’s function in regulating lipid homeostasis, which provides a mechanistic basis for ataxia that is distinct from peripheral neuropathy. Specifically, only ataxia-associated variants show enhanced lipid partitioning to mitochondria, uncoupling of mitochondrial fatty acid import from β-oxidation and enhanced ferroptotic vulnerability. Together, these findings identify lipid homeostasis as a previously underappreciated determinant of phenotypic diversity in MFN2 dysfunction and suggest that MFN2-linked ataxia arises through a specific pathogenic mechanism.

A central finding of this study is that cerebellar ataxia-associated MFN2 variants share a lipid droplet phenotype distinct from other MFN2 variants and MFN2 loss-of-function models. Previously, we observed a decrease in cellular lipid droplets in fibroblasts from a patient harbouring the D414V MFN2 variant (Sharma et al., 2022). However, this was a single patient, and we could not exclude whether other genetic changes might contribute to this observation. Here, using both multiple patient cells and isogenic cells re-expressing MFN2 variants, we see a consistent signature of reduced cellular lipid droplets that is only seen with MFN2 variants linked to cerebellar ataxia (T105M/T206I/R280C/D414V). This reduction in cellular lipid droplets contrasts with all of the other MFN2 variants in our panel, including those linked to CMT2A, which display an increased abundance of cellular lipid droplets, in accordance with previous reports (Hu et al., 2024; Larrea et al., 2019; Zaman et al., 2026). The consistency of the reduced lipid droplet phenotype across multiple ataxia variants, and its absence in other MFN2 variants, strongly argues that it is a conserved feature of MFN2-associated cerebellar ataxia.

The mechanisms underlying the multiple pathologies associated with MFN2 variants remain poorly understood. Our results here support the idea that MFN2-linked ataxia is due to a gain-of-function, specifically increased transfer of lipids from LDs to mitochondria. We propose this particular lipid perturbation is due to a gain-of-function as MFN2 has an established role in mediating LD-mitochondrial contacts (Boutant et al., 2017; Cai et al., 2024; Chung et al., 2019; Hu et al., 2024), and because we observe that MFN2 loss-of-function decreased mitochondrial lipid import and increased lipid droplet abundance, a finding consistent with previous reports (Bórquez et al., 2024; Cai et al., 2024). Intriguingly, the LD-mitochondrial gain-of-function we describe in ataxia-linked MFN2 variants was unexpected, as earlier work showed that cerebellum-specific MFN2 KO causes cerebellar degeneration (Chen et al., 2007). This KO work would suggest that cerebellar ataxia in patients with MFN2 variants could be caused by loss of MFN2 function. However, it is important to recognize that a knockout is much more severe than a pathogenic variant, in particular for a multifunctional protein such as MFN2. While the KO work shows that MFN2 is important for cerebellar function, it does not identify which MFN2 function(s) is/are critical, nor does it preclude the gain-of-function mechanism we describe here. Along these lines, the concepts of loss-of-function and gain-of-function variants become murky when a protein has multiple functions, as a variant can increase one function and impair another simultaneously. We posit that this complication contributes to the variable disease phenotypes associated with MFN2, with different mechanisms underlying certain MFN2 pathologies. Supporting this notion, we previously described a link between MFN2-linked myopathy and mtDNA-mediated inflammation (Zaman et al., 2025). Here, the fact that ataxia-linked variants produce a distinct gain-of-function lipid phenotype further helps to tease apart the mechanisms underlying MFN2 pathologies.

In addition to the altered lipid droplet phenotype, we observed a striking uncoupling of β-oxidation from fatty acid import in the ataxia MFN2 variants. Although the ataxia variants exhibited increased mitochondrial fatty acid import, they also displayed the lowest rates of β-oxidation among the variants examined. This reduced β-oxidation is evidenced both by reduced fatty acid oxidation observed with the FAOBlue dye and by the accumulation of acylcarnitine intermediates. These findings are consistent with previous studies showing that mitochondrial fatty acid overload can stall β-oxidation (Guerra et al., 2022; Saukko-Paavola & Klemm, 2024). Rather than coupling fatty acid import to efficient mitochondrial utilization, the ataxia-associated variants appear to disrupt this relationship, creating a cellular state in which fatty acids imported into the mitochondria are not efficiently broken down. Consistent with altered lipid transfer and inhibition of β-oxidation, we observed significant remodelling of the cellular lipidome. Major changes were seen in membrane phospholipids such as phosphatidylethanolamine, mitochondrial phospholipids such as cardiolipin, and other unsaturated lipid species prone to peroxidation.

The extensive remodelling of the lipid environment in ataxia-associated MFN2 variants raises the important question of how these changes contribute to disease pathogenesis. Increasing evidence links lipid peroxidation and ferroptosis to multiple forms of inherited ataxia. A key example is Friedreich’s ataxia, where mitochondrial dysfunction, oxidative stress and ferroptotic signalling are implicated in disease pathogenesis (Cotticelli et al., 2019; La Rosa et al., 2020; Turchi et al., 2020). Within this context, our findings point towards ferroptosis as a key downstream consequence of the altered lipid state observed in the ataxia-linked MFN2 variants. Across our KO-re-expression cells and the patient fibroblasts, the ferroptotic vulnerability is likely due to altered lipid dynamics. Notably, we did not observe substantial changes in cellular iron abundance or glutathione levels, which represent two common causes of ferroptotic susceptibility (Dar et al., 2026). Instead, our data support a model in which dysregulated lipid homeostasis contributes to ataxia by creating a lipid environment permissive for oxidative damage and ferroptosis.

Another key question that remains is how lipid alterations and vulnerability to ferroptosis disproportionately affect the cerebellum, leading to cerebellar ataxia. Growing evidence suggests lipid dyshomeostasis is a recurring feature across multiple inherited cerebellar ataxias, including disorders caused by pathogenic variants in genes such as *SNX14* (Thomas et al., 2014), *PNPLA6* (Wiethoff et al., 2017) and *ABHD13* (Joshi et al., 2018). Meanwhile, in mice, neuron-specific inactivation of GPX4 leads to cerebellar hypoplasia and loss of Purkinje cells, a key cerebellar cell type important for coordinating motor activity (Wirth et al., 2014). Given that GPX4 plays a vital role in combating ferroptosis, these findings support the notion that the cerebellum, and Purkinje cells specifically, are more susceptible to ferroptosis. Together, these observations suggest that the cerebellum is particularly sensitive to disruptions in lipid homeostasis and oxidative lipid damage, providing a potential explanation for why the lipid phenotypes associated with MFN2 variants manifest clinically as cerebellar ataxia.

Our findings also have potential therapeutic implications, as the robust cell death rescue with ferrostatin-1 suggests that ferroptosis may be a tractable target for MFN2-associated ataxia. Notably, current therapies for inherited cerebellar ataxias target pathways that are relevant to ferroptosis. For example, omaveloxolone activates NRF2-dependent antioxidant responses and has recently been approved for Friedreich’s ataxia (Pilotto et al., 2024). Likewise, the mitochondrial-targeted therapeutic elamipretide shows promise in reducing oxidative stress in Friedreich’s ataxia and preserving mitochondrial function by modulating cardiolipin (Pharaoh et al., 2023; Scott et al., 2024). As our findings suggest therapeutic benefit of limiting lipid peroxidation and ferroptosis, either omaveloxolone or elamipretide may be relevant treatments for MFN2-associated ataxia.

Finally, our findings broaden the emerging links between MFN2 and ferroptosis. Previous literature suggests that MFN2 is a positive regulator of ferroptosis as MFN2 overexpression induces ferroptosis (Wei et al., 2020), while MFN2 KO suppresses ferroptosis (Li et al., 2021; Sassano et al., 2025). In this latter example, mito-ER contact sites (MERCs), which are regulated by MFN2 (de Brito & Scorrano, 2008), are implicated as sites of lipid peroxidation that contribute to ferroptosis, such that MFN2 loss and subsequent reduction in MERCs is proposed to be protective against ferroptosis. However, our work here describes a new mechanism by which MFN2 promotes ferroptosis by altering lipid homeostasis via LD interactions with mitochondria. Notably, given that the D414V ataxia variant also perturbs MERCs (Sharma et al., 2022), it seems that in the context of ferroptosis, these LD perturbations can overcome any protective effects of reduced MERCs. Together, these studies highlight an important role for MFN2 in regulating ferroptosis through multiple mechanisms.

Despite the novel findings described here, some limitations should be considered. First, our isogenic KO-re-expression system models homozygous variant expression. Although the D414V variant represents a homozygous genotype, most MFN2 disorders are inherited in a heterozygous manner (Table 1). While the MFN2 KO background allows the effects of individual variants to be studied without the confounding effect from having a WT copy of MFN2, this approach also represents a more severe loss-of-function scenario than most patient fibroblasts. Importantly, we get consistent findings using either KO-re-expression lines or fibroblasts. Second, our findings are from a non-cerebellar cell culture system and therefore do not address the selective vulnerability of cerebellar neurons. While the evidence provided above argues strongly for a link between ferroptosis being a key driver of ataxia, future studies examining this pathway in cerebellar tissue would help support the argument.

Collectively, our findings support a model in which MFN2 dysfunction drives lipid dyshomeostasis and contributes to cerebellar ataxia, distinct from dysfunction underlying peripheral neuropathy and other MFN2 pathologies. By uncoupling mitochondrial fatty acid flux from downstream β-oxidation, ataxia variants promote pathogenic lipid remodelling, increasing susceptibility to oxidative damage and ferroptotic cell death. More broadly, these findings identify lipid homeostasis as an important determinant of phenotypic heterogeneity in MFN2 disease and suggest that ferroptosis represents a potential converging mechanism across multiple forms of inherited cerebellar ataxia.

## Data Availability

All data produced in the present work are contained in the manuscript

## Acknowledgements

The authors would like to thank the study participants and their families. We thank the Care4Rare Canada Consortium for access to cell line carrying the MFN2 R707W variant.

## Author contributions

Designed and/or performed and analyzed mitochondrial experiments: MZ, AM, CC, TS. Designed and/or performed and analyzed lipidomics experiments: MZ, AM, FSM, ABZ, TS. Clinical evaluation of patients and patient cells: GP. Drafted manuscript and figures: MZ, AM, TS. All authors discussed, commented on and consented authorship on the manuscript.

## Funding

This work was supported by funds provided by the Canadian Institutes of Health Research Project Grant (TES) and the Alberta Children’s Hospital Research Institute (Owerko Center) (MZ). MZ was supported by a Hotchkiss Brain Institute International Recruitment Scholarship and a University of Calgary Open Doctoral Scholarship. AM was supported by a Canadian Institutes of Health Research Canada Graduate Scholarship. The funders had no role in the study design, data collection and interpretation, or the decision to submit the work for publication.

**Extended Figure 1A:**
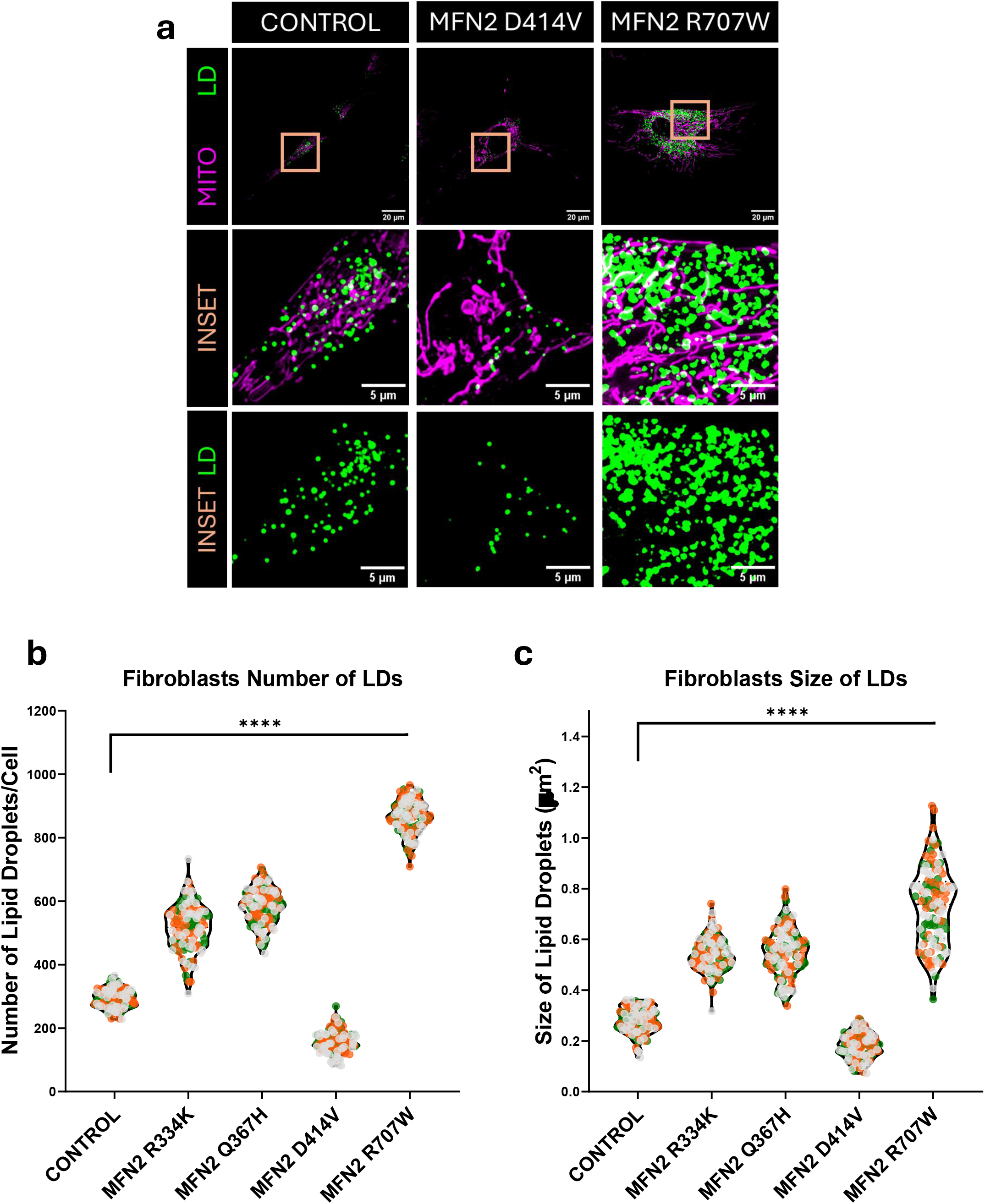
Ataxia-linked patient cells show reduced cellular lipid droplets. (a) Representative confocal live-cell images showing lipid droplets (LipidSpot 610) and mitochondria (MitoTracker Green) in healthy and patient fibroblasts (100x objective). The orange boxes in the top panels represent the area zoomed into in the bottom panel. (b-c) Quantitative analyses of the (b) number and (c) size of lipid droplets in patient fibroblasts. The violin plots show median and interquartile range; each dot represents a cell and colours represent biological replicates. The statistical symbols on the graphs indicate: **** P ≤ 0.0001.

**Extended Figure 1B:**
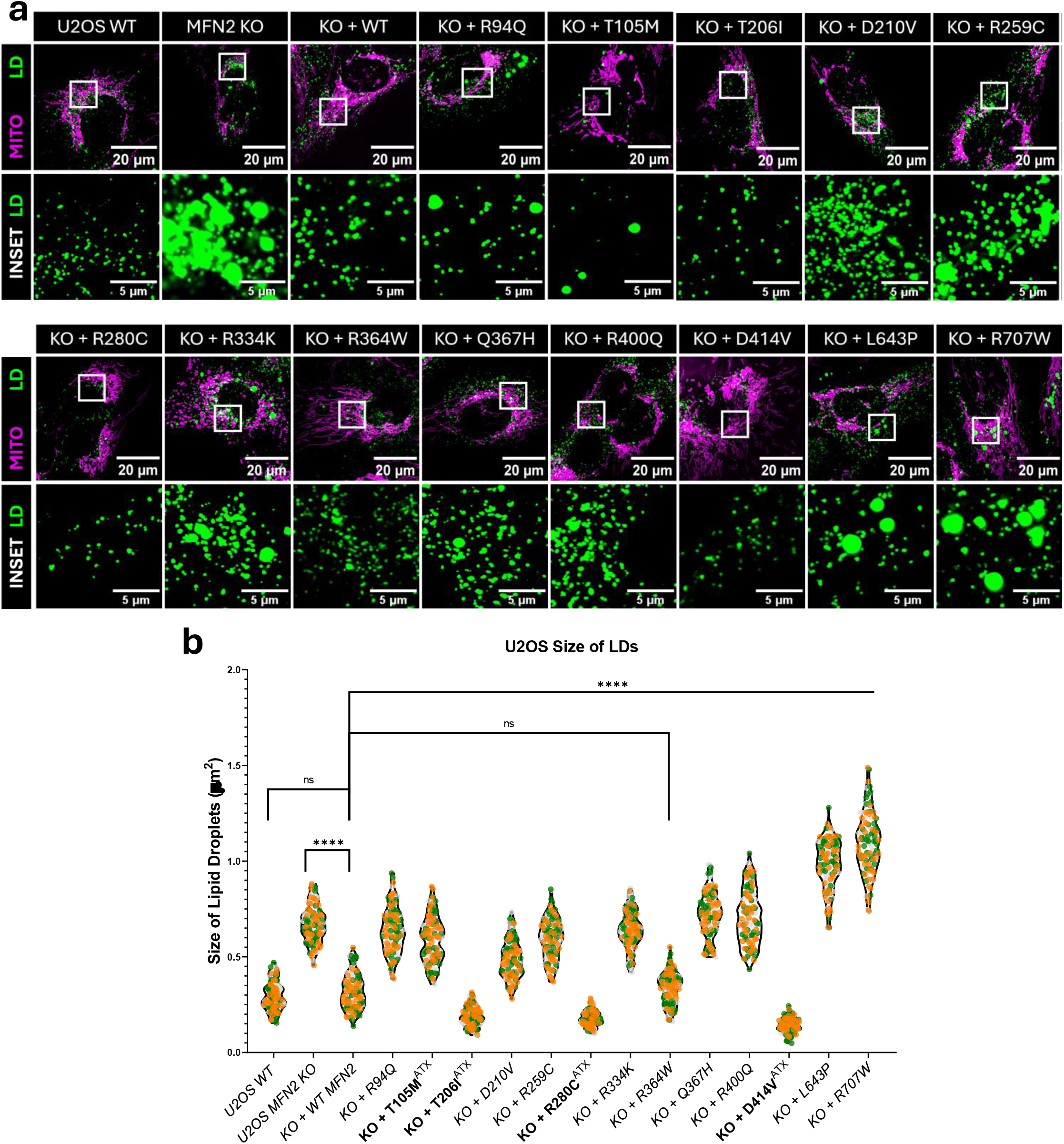
Ataxia-linked variants show reduced cellular lipid droplets. (a) Representative confocal live cell images (100x objective) showing lipid droplets (LipidSpot 610) and mitochondria (MitoTrackerGreen) in all the U2OS cell lines of interest. The white boxes in the top panels represent the area zoomed into in the bottom panel. (b) Quantitative analyses of the average size of lipid droplets per cell in U2OS cell lines of interest; the ataxia-specific variants are indicated on the graph. The violin plots show median and interquartile range, each dot represents a cell and colours represent biological replicates. The statistical symbols on the graphs indicate: ns P > 0.05, **** P ≤ 0.0001.

**Extended Figure 2:**
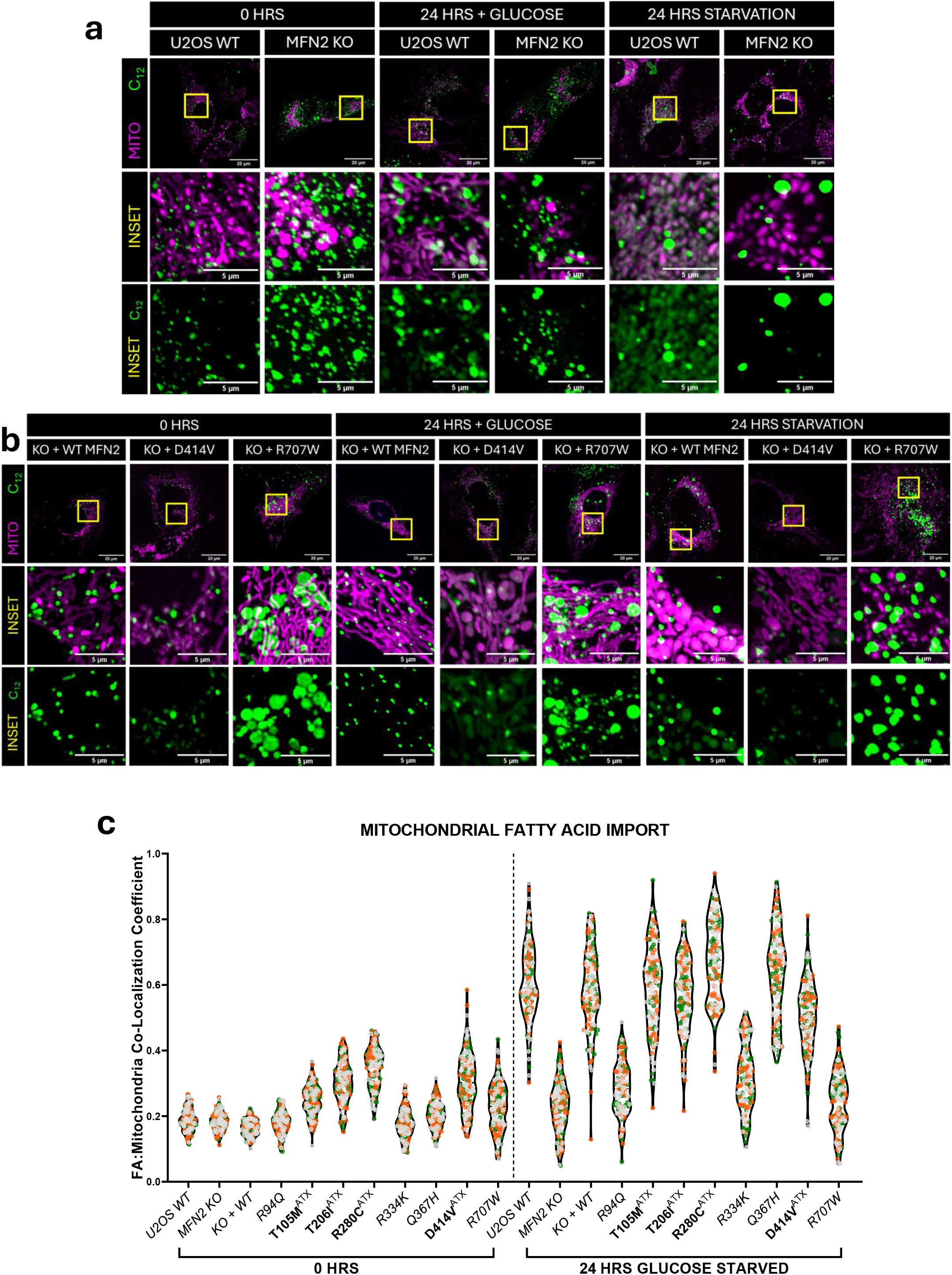
Ataxia-linked MFN2 variants show increased mitochondrial fatty acid import. (a-b) Representative confocal live cell images (60x objective) showing mitochondria (MitoTracker DeepRed) and fatty acids (BODIPY 558/568) in (a) U2OS WT/MFN2 KO cells and (b) WT re-expression/D414V re-expression/R707W re-expression after chasing with glucose supplementation for 24 hours. The white boxes in the top panels represent the area zoomed into in the bottom panel. (c) Quantitative analyses of Pearson Co-localization coefficient of mitochondria and BODIPY 558/568 fatty acids in U2OS cells at 0 hours and 24 hours chasing without glucose supplementation. The ataxia-specific variants are indicated on the graph. The violin plots in (c) and (e) show the median and interquartile range, each dot represents a cell and colours represent biological replicates.

**Extended Figure 3:**
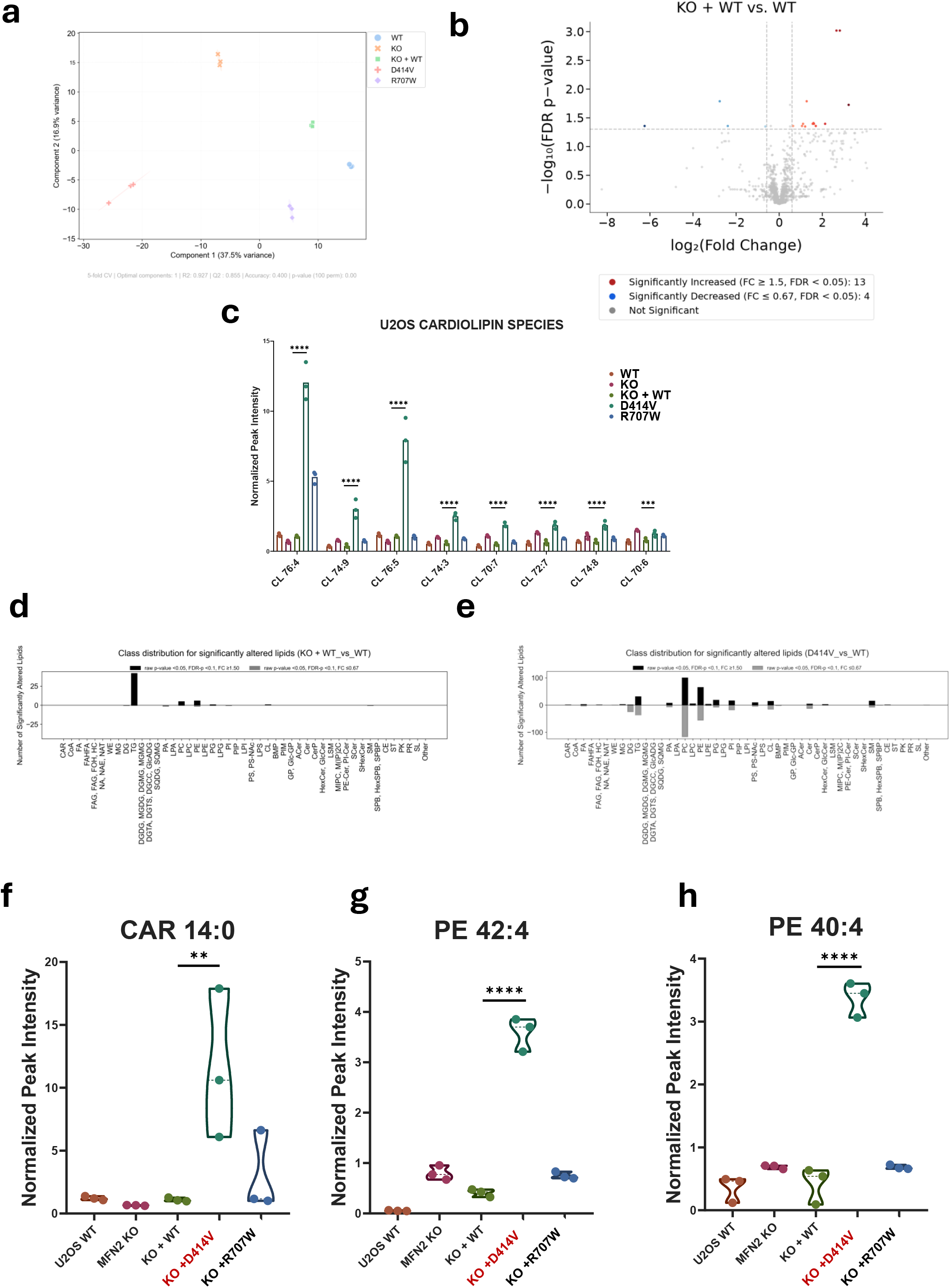
Remodelling of the lipidome in MFN2 D414V re-expression cells. (a) Principal component analysis of lipidomic analysis of U2OS cells. (b) Representative volcano plot comparing the lipidome in U2OS MFN2 KO cells re-expressing WT MFN2 vs WT U2OS cells. The top significant species are labelled on the plot. FC: fold-change for Group A / Group B. FDR-p: p-value corrected for false discovery rate (Benjamini-Hochberg correction). Lipids are considered significantly altered for FC ≥1.50 or ≤0.667, FDR-corrected p-value < 0.05. (c) Quantitative analysis of representative cardiolipin species in U2OS cells; the bars represent the mean values for each group. (d-e) Analysis of significantly upregulated lipid groups in (d) U2OS MFN2 KO cells re-expressing WT MFN2 vs U2OS WT cells and (e) U2OS MFN2 KO cells re-expressing MFN2 D414V vs re-expressing WT MFN2. (f-h) Quantitative analysis of levels of (f) acylcarnitine 14:0, (g) phosphatidylethanolamine 42:4 and (h) phosphatidylethanolamine 40:4 in U2OS cells. The violin plots in (f-h) represent the median and IQR of lipid species, n=3 biological replicates.

**Extended Figure 5:**
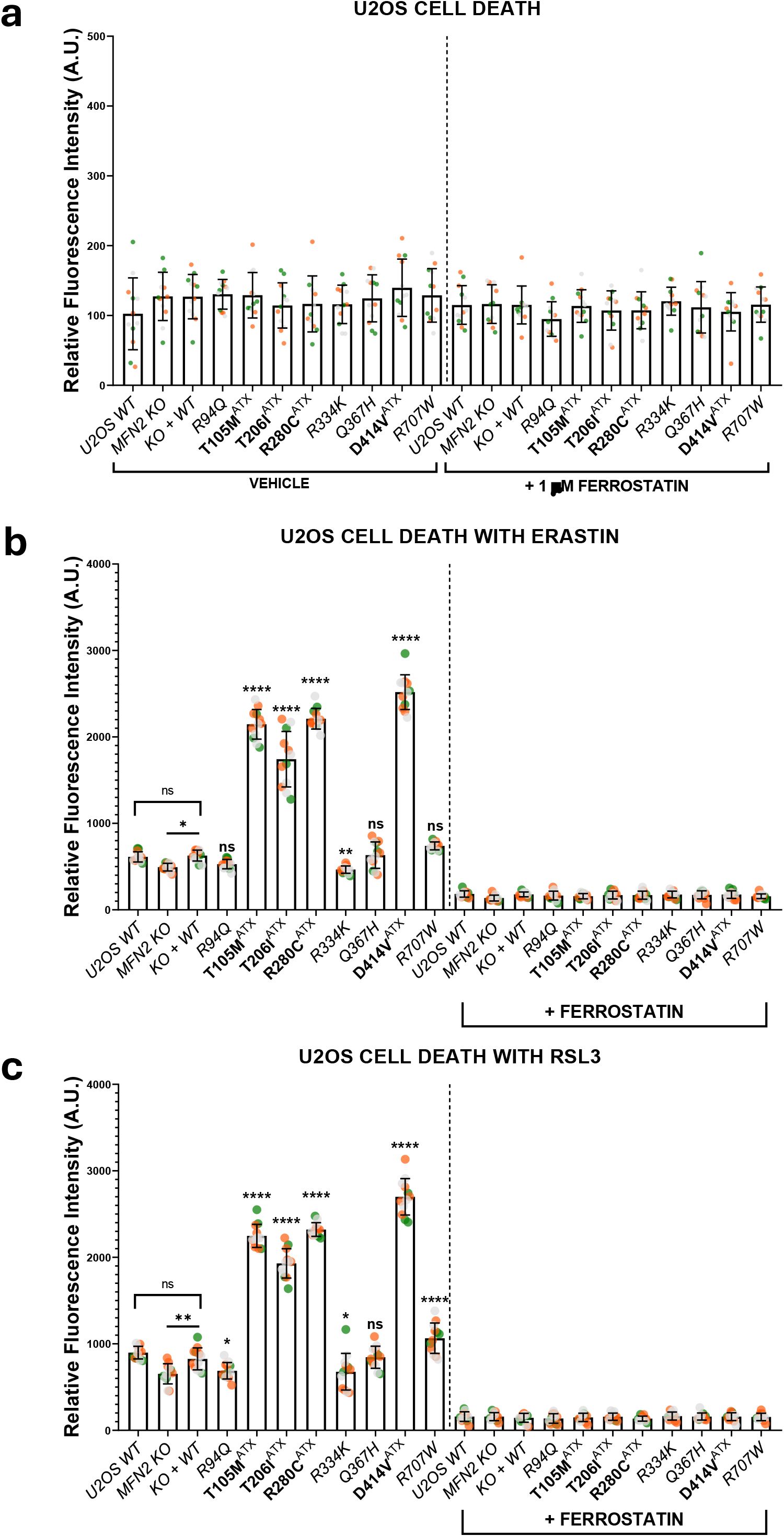
Increased ferroptotic cell death in ataxia-linked variants. (a) Quantification of fluorescent intensity from Sytox Orange dye indicating cell death in U2OS cells, untreated or treated with 1uM ferrostatin. (b-c) Quantitative analyses of fluorescent intensity from Sytox Orange dye indicating cell death in U2OS cells, treated with (b) 5uM Erastin or 5uM Erastin and 1uM ferrostatin and (c) 1uM RSL3 or 1uM RSL3 and 1uM ferrostatin. The bars and lines in (a-c) indicate mean +/-SD, each point indicates a well and colours represent biological replicates. The statistical symbols on the graphs indicate: ns P > 0.05, * P ≤ 0.05, ** P ≤ 0.01, **** P ≤ 0.0001.

**Extended Figure 6:**
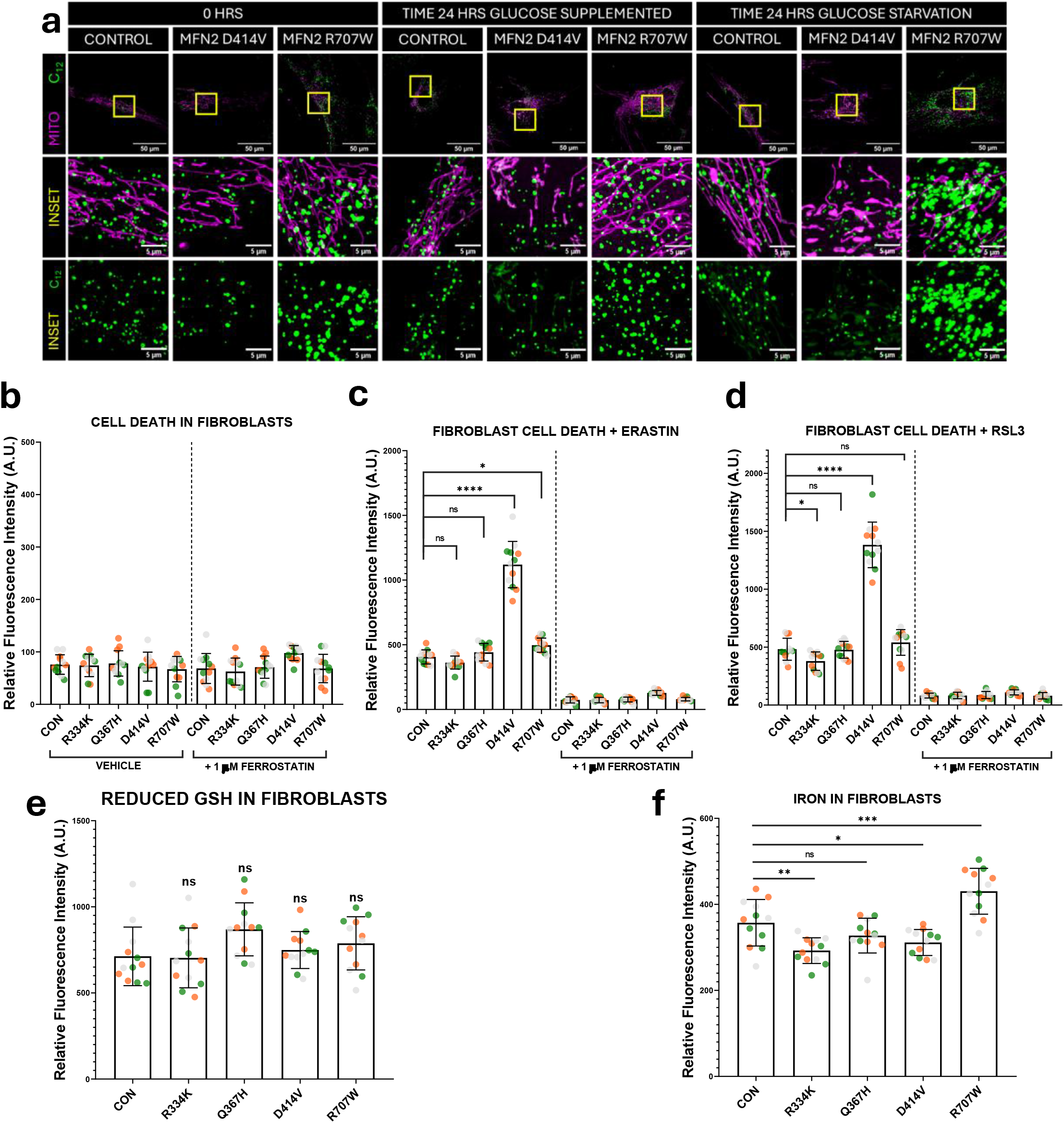
Ataxia-linked fibroblast replicates ferroptotic phenotypes. (a) Representative confocal live cell images (60x objective) showing mitochondria (MitoTrackerGreen) and fatty acids (BODIPY 558/568) in healthy and MFN2 patient fibroblasts at 0 hours and at 24 hours, supplemented with or without glucose. The yellow boxes in the top panels represent the area zoomed into in the bottom panel. (b) Quantification of fluorescent intensity from Sytox Orange dye indicating cell death in fibroblasts, untreated or treated with 1uM ferrostatin. (c-d) Quantitative analyses of fluorescent intensity from Sytox Orange dye indicating cell death in fibroblasts, treated with (c) 5µM Erastin or 5uM Erastin and 1uM ferrostatin and (d) 1µM RSL3 or 1µM RSL3 and 1µM ferrostatin. (e-f) Quantitative analyses of (e) levels of reduced GSH, (f) levels of iron in fibroblasts. The bars and lines in (b-f) indicate mean +/-SD, each point indicates a well and colours represent biological replicates. The statistical symbols on the graphs indicate: ns P > 0.05, * P ≤ 0.05, ** P ≤ 0.01, *** P ≤ 0.001, **** P ≤ 0.0001.

**Extended Figure 6B:**
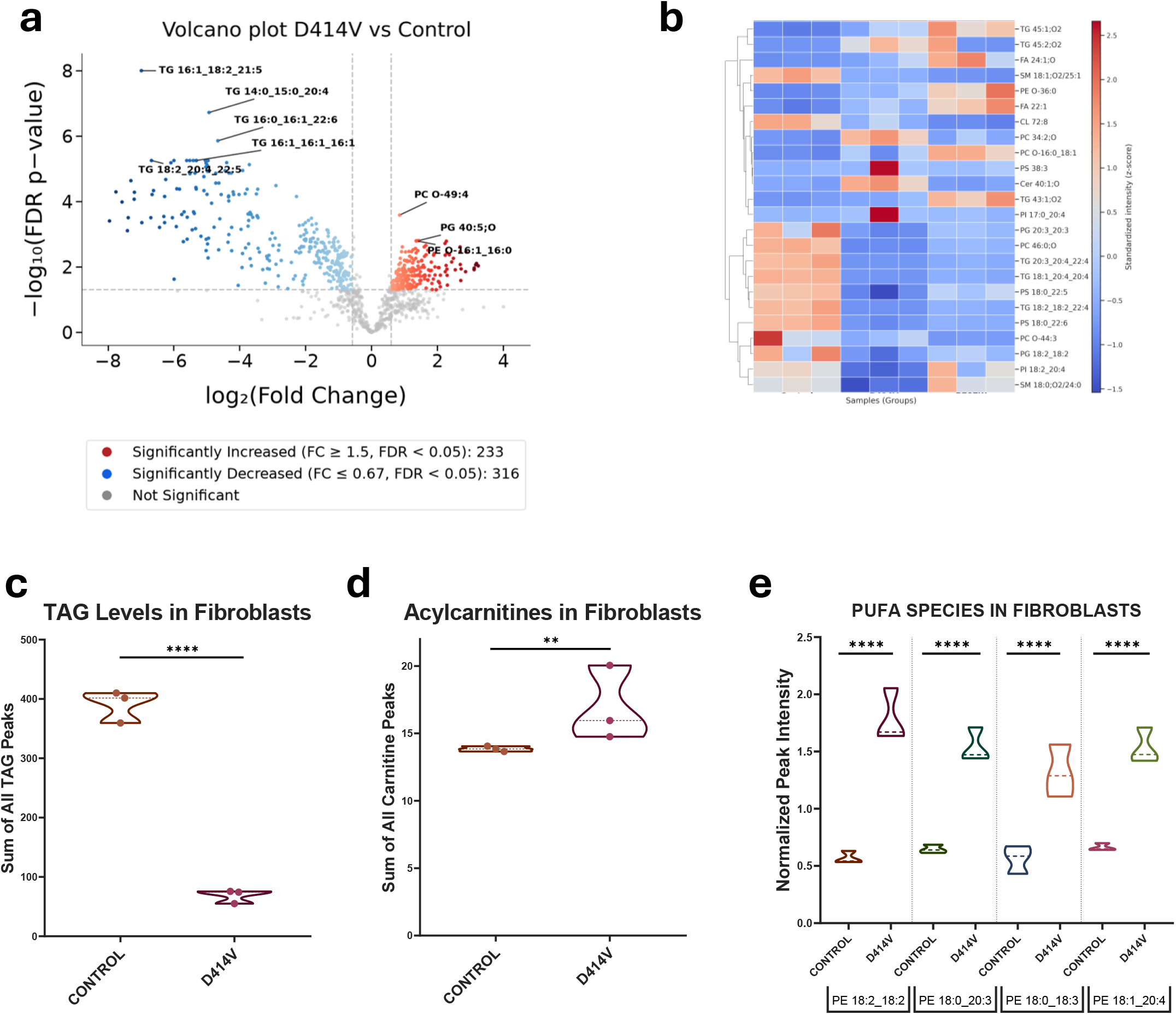
Ataxia-linked fibroblast replicates ferroptotic phenotypes. (a) Representative volcano plot comparing lipidome in D414V patient fibroblasts versus healthy control fibroblasts. (b) Representative heatmap showing top 25 lipid species altered in D414V fibroblasts. The top significant species are labelled on the plot. FC: fold-change for Group A / Group B. FDR-p: p-value corrected for false discovery rate (Benjamini-Hochberg correction). Lipids are considered significantly altered for FC ≥1.50 or ≤0.667, FDR-corrected p-value < 0.05. (c-d) Quantification of the sum of all (c) TAG species and (d) Acylcarnitine species in patient and control fibroblasts. (e) Quantitative analysis of the top four PUFA phosphatidylethanolamine species in patient and healthy fibroblasts. The violin plots in (c-e) represent the median and interquartile ranges, n=3 biological replicates.

